# Syndromic surveillance insights from a symptom assessment app before and during COVID-19 measures in Germany and the United Kingdom: results from repeated cross-sectional analyses

**DOI:** 10.1101/2020.06.16.20126466

**Authors:** Alicia Mehl, Francois Bergey, Caoimhe Cawley, Andreas Gilsdorf

## Abstract

**Background:** Unprecedented lockdown measures have been introduced in countries across the world to mitigate the spread and consequences of COVID-19. While attention has focused on the effects of these measures on epidemiological indicators relating directly to the infection, there is increased recognition of their broader health implications. However, assessing these implications in real time is a challenge, due to limitations of existing syndromic surveillance data and tools.

**Objective:** To explore the added value of mobile phone app-based symptom assessment tools as real time health insight providers to inform public health policy makers.

**Methods:** A comparative and descriptive analysis of the proportion of all self-reported symptoms entered by users during an Ada assessment in Germany and the United Kingdom (UK) was conducted between two periods: before and after the implementation of “Phase One” COVID-19 measures. Additional analyses were performed to explore the association between symptom trends and seasonality, and symptom trends and weather. Differences in the proportion of unique symptoms between the periods were analysed using Pearson’s Chi-squared test and reported as Log2 Fold Changes (Log2 FC).

**Results:** Between 48,300-54,900 symptomatic users reported 140,500-170,400 symptoms during the Baseline and Measures periods in Germany. Between 34,200-37,400 symptomatic users in the UK reported 112,100-131,900 symptoms during the Baseline and Measures periods. The majority of symptomatic users were female (Germany 68,600/103,200, 66.52%; UK 51,200/71,600, 72.74%). The majority (Germany 68,500/100,000, 68.45%; UK 50,900/68,800, 73.91%) were aged between 10 and 29 years, and about a quarter (Germany 26,200/100,000, 26.15%; UK 14,900/68,800, 21.65%) were between 30-59 years. 103 symptoms were reported either more or less frequently (with statistically significant differences) during the Measures as compared to the Baseline period, and 34 of these were found in both countries. The following mental health symptoms (Log2 FC, *P*-value) were reported less often during the Measures period: *inability to manage constant stress and demands at work* (−1.07, *P*<.001), *memory difficulty* (−0.56, *P*<.001), *depressed mood* (−0.42, P<.001), and *impaired concentration* (−0.46, *P*<.001). *Diminished sense of taste* (2.26, *P*<.001) and *hyposmia* (2.20, *P*<.001) were reported more frequently during the Measures period. None of the 34 symptoms were found to be different between the same dates in 2019. Fourteen of the 34 symptoms had statistically significant associations with weather variables.

**Conclusions:** Symptom assessment apps have an important role to play in facilitating improved understanding of the implications of public health policies such as COVID-19 lockdown measures. Not only do they provide the means to complement and cross-validate hypotheses based on data collected through more traditional channels, they can also generate novel insights through a real-time syndromic surveillance system.

## Introduction

### Background

Since its emergence at the end of 2019, COVID-19 has had an enormous and wide-ranging impact, with millions of confirmed cases and hundreds of thousands of deaths reported worldwide [1]. Governments have introduced a series of measures ranging from work and school closures to social distancing and lockdowns to mitigate the spread and consequences of infection. The measures have been unprecedented on many levels: in how disruptive they are to daily life, by the proportion of populations affected, in the duration of implementation, and by their global reach. As a result, the daily lives of millions have been upended.

The extent of the counter-measures to the pandemic raises questions as to their impact on public and individual health. While much of the initial focus in the medical and scientific community has been on understanding the virus and infection itself [2], as well as on the effects of various policy measures on epidemiological indicators relating directly to COVID-19 [3,4], there has also been increased recognition of the broader health effects [5].

Some of the broader implications concern the direct and immediate consequences of lockdown. For instance, as people are distressed due to social isolation and the economic fallout of the crises, an upsurge in the incidence and severity of mental health problems has been predicted [5–11]. Increased handwashing (a primary recommendation to reduce transmission of COVID-19) is expected to result in increased skin irritation and dermatitis [12,13]. Other implications are related to more indirect factors such as delays and cancellations to surgeries and non-urgent treatments for cancer and other patients [14–16], interruptions in drug and commodities supply chains [17,18], and drops in vaccination rates for vaccine-preventable diseases [19,20]. The consequences of these interruptions to medical services during this pandemic will likely create a higher morbidity, but the impact may not be visible for several years to come [17].

Assessing the health implications of lockdown policies in real-time is a challenge. Apart from the time lag, evidence cited in support of purported health effects is often anecdotal or based on surveys conducted among medical professionals. The latter provide important insights into changes in health symptoms identified at the point of contact, but may not account for the effects of social distancing guidelines on health-seeking behavior. Traditional syndromic surveillance data [21] provides valuable information for a defined set of indicators over time, but is unable to distinguish whether the trends reflect changes in disease incidence or in the uptake of health services. More significant efforts towards data collection among the general public are currently being undertaken [22]. However, these are missing a Baseline prior to the emergence of COVID-19 that would facilitate a clearer understanding of the impact. In general, surveys can provide snapshots of a highly dynamic situation, but tend to be restricted in scope, meaning that changes in health beyond pre-defined indicators risk being overlooked.

In the past years, a growing number of studies have shown the benefits of mobile phone app-based symptom assessment tools for improved health outcomes, for example, in lowering the barrier to seek help [23–29]. If the literature on symptom assessment tools has primarily focused on individual health benefits, e.g. early detection of rare diseases [30], recent research has also demonstrated their potential contribution to public health. Specifically with regard to COVID-19, symptom tracker apps have helped flag anosmia (loss of the sense of smell) as a potentially relevant symptom of infection [31,32], as well as to challenge common understandings of presenting symptoms of COVID-19 [33].

### Goal of this study

In the present study, we add to this body of literature by exploring the added value of symptom assessment tools for the analysis of broader health implications resulting from public health interventions such as lockdown measures. The study is based on self-reported symptom data from Ada, a digital symptom assessment app which uses a probabilistic reasoning engine that collects demographic information, symptoms, and medical history to suggest possible conditions and then guide individuals to the most appropriate care. The Ada app is available in seven languages and has over ten million users. It is described in Gilbert, et al in further detail [34]. Using an inductive approach and focusing on the immediate impact of COVID-19 control measures, we compared symptom data reported by users in Germany and the UK before and during the first phase of COVID-19 lockdown measures to identify changes and continuities in the incidence of self-reported symptoms.

The aim of this study was to explore the potential of the Ada symptom assessment app to generate real-time health insights to inform public health policy makers.

## Methods

### Study focus

This analysis is a comparative descriptive study of self-reported symptoms entered by users in an Ada assessment completed during the time of the “Phase One” COVID-19 interventions (the Measures period) compared to a Baseline period in Germany and the United Kingdom. The Measures periods began in each country when all five major non-pharmaceutical interventions described in Flaxman et al [4] were implemented (specifically: school closure ordered, case-based measures (strong recommendation of self-isolation when showing COVID-19-like symptoms), public events banned, social distancing encouraged, lockdown decreed) and lasted until April 22nd, 2020, when data were extracted. The end of the Baseline period was defined as the day before any of the five interventions were implemented in the respective country, and the length of the Baseline period was equal to the Measures period in the same country. The period between the Baseline and Measures periods is excluded from the analysis due to the partial implementation of “Phase One” COVID-19 measures. This is an exploratory analysis of all symptoms (not only COVID-related) reported by symptomatic users during the defined periods. In this study, we consider a symptomatic user as one who completed at least one assessment during the periods of analysis, either themselves or by someone on their behalf (i.e. a legal guardian if under 16 years). We considered trends if in both countries, the proportion that a specific symptom was reported (out of all reported symptoms) was significantly different between the two periods.

### Additional analyses

Ad-hoc analyses were later performed to test identified trends for symptoms against selected potential confounders, such as seasonality and weather. To explore the potential impact of seasonality on trends, the results were compared to the same analysis conducted for the same dates in 2019. To explore the impact of weather on trends, associations between the monthly proportion of reported symptoms and weather variables (average monthly temperature, monthly precipitation (mm), and monthly hours of sunshine) were investigated during the period January 2019 - March 2020. The weather analysis was restricted to Germany.

### Participants

All assessments completed by Ada users in Germany and the UK during the Baseline and Measures period were included in the analysis. We analysed pseudonymized health data for public health purposes, according to the European General Data Protection Regulation (GDPR), and users are duly informed of such use of their data (information available at any time in Ada’s Privacy Policy). Additionally, users maintain their right to object to such processing for reasons arising from their particular situation, as required by the GDPR.

### Variables

An Ada assessment consists of different parts: 1) the user enters an unlimited number of symptoms, 2) the user is asked about other potential symptoms they could have, and 3) an assessment result is provided with conditions that could potentially explain the reported symptoms and adequate triage. This analysis only includes symptoms that are self-reported by a user in the first part of the assessment: that is, responses to the initial question “*Let’s start with the symptom that’s troubling you the most*”, followed by “*Do you have any other symptoms?*”. Upon entering free text, the user is then given a range of medically-curated options (based on linguistic relevance) to select from.

The variable of interest *S*_*i,k*_, representing the proportion a symptom *i* is self-reported during the period *k* is defined as

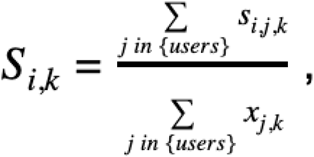

where

*s*_*i,j,k*_ equals 1 if the user *j* self-reported the symptom *i* during the period *k*, and equals 0 otherwise.

*x*_*j,k*_ equals 1 if the user *j* has finished one assessment during the period *k*, and equals 0 otherwise.

The age variable was grouped using the following categories (years): 0-9, 10-19, 20-29, 30-39, 40-59 and 60+.

Weather data (average monthly temperature (°C), monthly precipitation (mm) and monthly hours of sunshine) for Germany were extracted from the Deutscher Wetter Dienst database for the period January 2019 until March 2020 [35].

For ease of reporting and to aid interpretation, symptoms which were reported in significantly different proportions during the Baseline and Measures periods were grouped using the International Classification of Diseases version 2019 (ICD-10) of the World Health Organization [36]. ICD-10 R sub-groups, named “Symptoms, signs and abnormal clinical and laboratory symptoms, not elsewhere classified” were used when possible. Similar categories were grouped together later, as presented in (Table 1).

**Table 1.** List of groups based on the ICD-10 classification.

|  |  | ICD-10 classification |
| --- | --- | --- |
| **Group name** |  |  |
|  | Circulatory and respiratory systems | R00- R09, J |
|  | Digestive system and abdomen | R10 – R19 |
|  | Skin and subcutaneous tissue | R20 – R23, L |
|  | Musculoskeletal | M |
|  | Genitourinary system | R30 – R39, N |
|  | Cognition, perception, emotional state and behaviour | R40 – R46, F |
|  | Speech and voice | R47 – R49 |
|  | General symptoms and signs | R50 – R69 |
|  | Nervous system | G |
|  | Eye and adnexa | H |

### Bias

As Ada’s medical model and databases are continuously updated, we defined the Baseline period to be as close as possible to the Measures period to limit the impact of these changes on the data. All modeled symptoms that were added, deleted, significantly modified, or significantly affected by the modification of any other symptom from the first day of the Baseline period until the end of the Measures period were removed from this study.

### Statistical methods

Sex and age groups of symptomatic users were reported as percentages and tested for differences between the periods with Pearson’s Chi-squared test. Differences in the proportion of symptoms between the periods were reported as Log2 Fold Changes and were analyzed with Pearson’s Chi-squared test. A Log2 Fold Change of 0.5 means that the proportion of that reported symptom was 1.41 times as large during the Measures compared to the Baseline. A Log2 Fold Change of 1 is interpreted as being twice as large during the Measures compared to the Baseline, and a Log2 Fold Change of 2 is four times as large. Conversely, a Log2 Fold Change of -1 means that the proportion of the reported symptom was twice as large during the Baseline compared to the Measures period. In general, Log2 Fold Change calculations are helpful in understanding relative differences in the proportions of users reporting each symptom between the two periods, but do not reflect how common reporting of that symptom was overall. Associations between weather variables and the proportion of symptoms were tested based on the Spearman’s Rank correlation coefficient.

When required, *P*-values were adjusted for multiple testing using the false discovery rate method. *P*-values less than or equal to 0.05 were considered statistically significant. Statistical analyses and figures were executed using R (version 3.6.1).

The analysis was done using exact numbers, but results representing user numbers are presented rounded to the closest hundred, to ensure a fully anonymised presentation of the results.

## Results

An overview of the Baseline and Measures periods in Germany and the UK (numbers of Ada users, numbers of symptoms reported) are shown in (Table 2).

**Table 2.** Key parameters.

|  | Germany |  | United Kingdom |  |
| --- | --- | --- | --- | --- |
|  | Baseline | Measures | Baseline | Measures |
| Dates of period (dd.mm.yy) | 03.02.20 – 05.03.20 | 22.03.20 – 22.04.20 | 11.02.20 – 11.03.20 | 24.03.20 – 22.04.20 |
| Number of days per period | 32 | 32 | 30 | 30 |
| Number of all users (N<sub>a</sub>) | 467,000 | 483,100 | 488,800 | 501,300 |
| Number of symptomatic users (N<sub>a</sub>) | 54,900 | 48,300 | 37,400 | 34,200 |
| Number of reported symptoms by symptomatic users (N<sub>a</sub>) | 170,400 | 140,500 | 131,900 | 112,100 |
| Number of symptoms reported at least once in Baseline or Measures period (N<sub>a</sub>) | 1,328 |  | 1,294 |  |

<sup>a</sup> rounded to the nearest hundred

Demographic characteristics of users are shown in (Table 3). During both the Baseline and Measures periods, the majority (Germany Baseline: 36,300/54,900, 66.19%; Germany Measures: 32,300/48,300, 66.90%; UK Baseline: 26,600/37,400, 71.17%; UK Measures: 24,600/34,200, 71.94%) of symptomatic users in both countries were female. The majority were aged between 10 and 29 years (Germany Baseline: 37,000/53,200, 69.51%; Germany Measures: 31,400/46,800, 67.13%; UK Baseline: 27,200/35,800, 75.76%; UK Measures: 23,700/32,900, 71.89%). Those aged between 30-59 years represented roughly a quarter (Germany Baseline: 13,300/53,200, 24.94%; Germany Measures: 12,900/46,800, 27.53%; UK Baseline: 7,100/35,800, 19.92%; UK Measures: 7,700/32,900, 23.54%) of symptomatic users. The number of symptomatic users in the Baseline (54,900 Germany, 37,400 UK) was slightly higher than in the Measures period (48,300 Germany, 34,200 UK).

**Table 3.** Demographic characteristics of the study population.

|  |  | Germany |  |  |  |  | United Kingdom |  |  |  |  |
| --- | --- | --- | --- | --- | --- | --- | --- | --- | --- | --- | --- |
|  |  | Baseline |  | Measures |  | P-value<sup>c</sup> | Baseline |  | Measures |  | P-value<sup>c</sup> |
|  |  | N<sub>a</sub> | % | N<sub>a</sub> | % |  | N<sub>a</sub> | % | N<sub>a</sub> | % |  |
| **Sex** |  |  |  |  |  |  |  |  |  |  |  |
|  | female | 36,300 | 66.2 | 32,300 | 66.9 | 0.02 | 26,600 | 72.2 | 24,600 | 72.9 | 0.02 |
| **Age<sup>b</sup>**<br>**(years)** |  |  |  |  |  |  |  |  |  |  |  |
|  | 0-9 | 1,300 | 2.4 | 700 | 1.6 | <.001 | 900 | 2.4 | 700 | 2.0 | <.001 |
|  | 10-19 | 17,100 | 32.1 | 13,700 | 29.2 | <.001 | 16,200 | 45.1 | 13,600 | 41.2 | <.001 |
|  | 20-29 | 20,000 | 37.5 | 17,800 | 37.9 | 0.28 | 11,000 | 30.7 | 10,100 | 30.7 | 0.95 |
|  | 30-39 | 6,400 | 12.0 | 6,000 | 12.9 | <.001 | 3,100 | 8.7 | 3,300 | 9.9 | <.001 |
|  | 40-59 | 6,900 | 13.0 | 6,800 | 14.6 | <.001 | 4,000 | 11.2 | 4,500 | 13.6 | <.001 |
|  | 60+ | 1,700 | 3.2 | 1,800 | 3.9 | <.001 | 700 | 2.1 | 900 | 2.7 | <.001 |

<sup>a</sup> rounded to the nearest hundred
<sup>b</sup> Users who did not report a birth year were excluded from the analysis
<sup>c</sup> Pearson's chi-squared test for differences between Baseline and Measures

Twenty-one symptoms were excluded from the analysis as they had been added to or deleted from the medical model during either the Baseline or Measures period. Three additional symptoms were excluded from the analysis as there were significant changes in the associated terms (text entered by users to match the description of a symptom to the term used in the model), which could affect the number of times a symptom is reported. A list of these symptoms is included in Table A of the Supplementary Materials.

### Main analyses

Out of 1,328 and 1,294 symptoms investigated respectively in Germany and in the United Kingdom, 103 symptoms were reported either more or less frequently, in either country, during the Measures as compared to the Baseline period. The complete results can be found in (Tables B and C) of the Supplementary Materials.

Figure 2 presents the 34 symptoms that showed a statistically significant difference in both countries. 24 symptoms were reported less often and 10 were reported more often in the Measures period in comparison to the Baseline period.

**Figure 2.**
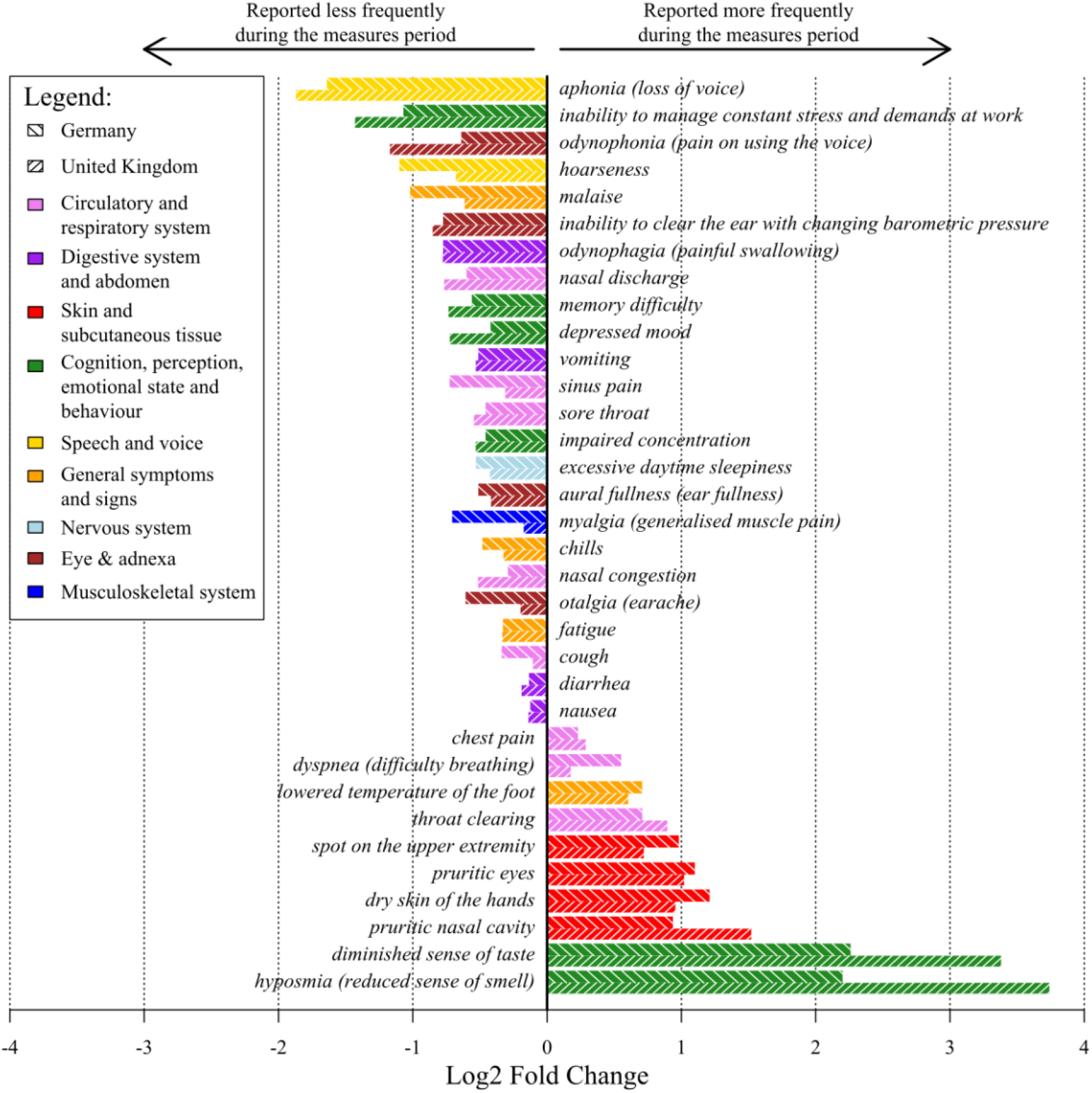
Relative difference (Log2 Fold Change values) in the proportions of Ada users’ reported symptoms with statistically significant differences between the Baseline and COVID-19 Measures periods in Germany and the UK

Out of the 34 significant symptoms, all skin and tissue-related symptoms (*pruritic nasal cavity, spot on the upper extremity, dry skin of the hands*, and *pruritic eyes*) were reported more frequently during the Measures period. In contrast, all speech and voice symptoms (*aphonia* and *hoarseness*), all eye and adnexa symptoms (*odynophonia, inability to clear the ear with changing barometric pressure, aural fullness*, and *otalgia*), all digestive system and abdomen symptoms (*odynophagia, diarrhea*, and *nausea*) and all musculoskeletal and nervous systems symptoms (*excessive daytime sleepiness* and *myalgia*) were reported less frequently during the Measures period.

In the cognition, perception, emotional state and behaviour symptoms group, two perception symptoms (*diminished sense of taste* and *hyposmia*), were reported more frequently during the Measures period whereas the mental health symptoms (*inability to manage constant stress and demands at work, memory difficulty, depressed mood*, and *impaired concentration*) were reported less frequently during the Measures period. Out of the circulatory and respiratory symptoms, three were reported more frequently during the Measures period (*throat clearing, dyspnea*. and *chest pain*) and four were reported less frequently (*nasal discharge, sinus pain, sore throat*, and *cough*). One general symptom was reported more frequently (*lowered temperature of the foot*) and three (*malaise, chills*, and *fatigue*) were reported less frequently during the Measures period.

### Additional analyses

Out of the 34 symptoms found to be different between the Baseline and Measures period in both Germany and the UK in 2020, none were found to be different between the same periods in 2019 in both countries. However, looking at the countries separately, in Germany, the data shows that eight of the 34 significant symptoms were also reported less frequently during the Measures period in 2019: *cough, chills, nasal discharge, myalgia, sore throat, malaise, fatigue* and *sinus pain*. The complete results can be found in Tables D and E of the Supplementary Materials.

The monthly weather reports in February 2020 (which corresponds to most of the Baseline period) differed in comparison to March 2020 (which corresponds to most of the Measures period) in Germany. Average temperature increased by 5.1°C (from 5.3°C to 10.4°C), monthly precipitation decreased by 107.8mm (from 124.1mm to 16.3mm) and the number of hours of sunshine per month increased by 228.5h (from 63.9h to 292.4h). Fourteen of the 34 significant symptoms had statistically significant associations with weather. Increased temperature was positively associated with *spot on the upper extremity* and negatively associated with *chest pain, lowered temperature of the foot, odynophagia, malaise, myalgia, cough, otalgia, chills, vomiting, sinus pain*, and *nasal congestion*. Increased hours of sunshine was positively associated with *pruritic eyes, spot on the upper extremity*, and *pruritic nasal cavity* and negatively associated with *lowered temperature of the foot* and *sinus pain*. The complete results can be found in Table F & Figure G of the Supplementary Materials.

## Discussion

### Principal Results

The results presented above show significant differences in the frequency and proportion of self-reported symptoms in Ada assessments before and after the implementation of measures aimed at reducing the transmission of COVID-19. Importantly, the same differences were found in both Germany and the UK, despite the divergent trajectories of these countries in the lead-up to the implementation of lockdown policies [37], as well as other national differences such as those relating to health systems [38]. Furthermore, these differences were not found during the same time periods in 2019, suggesting that the lockdown measures could have contributed to the results.

Many of the observed differences were to be expected. The reduced frequency of reported respiratory symptoms (*nasal discharge, sore throat, cough, sinus pain, nasal congestion, hoarseness, odynophonia, aphonia*) and influenza-like illness (i.e. *malaise, fatigue, chills, myalgia*) following the measures is understandable as the cold and flu season has also waned exceptionally rapidly during this period [39], likely facilitated by reduced contact resulting from lockdown. To support this interpretation, *cough, chills, nasal discharge, myalgia, sore throat, malaise, fatigue* and *sinus pain* were also reported less frequently during the Measures periods in 2019 in Germany, suggesting that seasonal changes are reflected in the data. The increased reporting of dry hands following the measures is consistent with more frequent handwashing during the Measures period, as expected by dermatologists [12,13]. Increased reports of *pruritic eyes* and *pruritic nasal cavity* following the measures could be a consequence of seasonal hay fever (known to be worse during the Spring compared to Winter months due to increased pollen in the air [40]), as these symptoms were found to be associated with increased sunshine (presumably when people spend more time outdoors).

The reduction of gastrointestinal symptoms could be associated with the closing of preschool/ day care settings (as they are known to contribute to the spread of these diseases [41]), restaurant closures, or due to improved hand hygiene. The decrease of ear problems could be a result of the extremely reduced air travel with much less people experiencing pressure adjustment problems called “airplane ear” [42], or related to the end of the cold and flu season. In these cases more research would be needed to explore these hypotheses.

We have also observed an increase in the reporting of *hyposmia, diminished sense of taste*, and *dyspnea* during the Measures period. These are less frequent but also typical COVID-19 symptoms and were increasingly recognized in the general public [31,43,44]. They were not found to be associated with seasonality or weather. This increase could be related to COVID-19 infections, or an artefact resulting from increased awareness of these symptoms due to media coverage.

A more surprising result is that *depressed mood, inability to manage constant stress and demands at work, impaired concentration, memory difficulty*, and *excessive daytime sleepiness* were reported in a lower proportion during the Measures period. This is not only contrary to conventional wisdom, but also runs counter to what was observed during previous infectious disease outbreaks, such as SARS, MERS, and Ebola [45,46] as well as to the literature reporting on the effects of COVID-19 on Mental Health [6–11,47,48]. Despite the fact that during the Measures period the temperature was warmer and there was more sunshine than usual, the analysis on weather data for Germany did not show a significant impact on the changed mental health symptoms. Our findings are supported by a growing body of evidence from ongoing studies [49] and recently published research [50], as well as anecdotal evidence reported in various mainstream media reports [51-53] which suggest that at least in the short-term the mental health effects of the COVID-19 measures may not be as negative as expected. One factor may be that the stress of everyday life and work/study is reduced during lockdown and home office/schooling. In addition, the reduction of *excessive daytime sleepiness* may be due to the fact that people may sleep better or more during lockdown as there are less available places to go out and socialize in the evenings. Ongoing studies into the mental health effects of COVID-19 and its countermeasures may shed further light on these questions.

### Limitations

It is important to interpret the study results taking into consideration the characteristics of the study population and the normal use case for Ada. Due to the specific age and sex distribution of users (predominantly young and female), the results may not be generalisable to other population groups. Furthermore, this analysis was limited to Germany and the UK (due to sufficient user numbers), and represents a two month snapshot of reported symptoms. Users who know they have a disease might not use Ada if their symptoms deteriorate, as the cause is already known. In addition, this is an analysis of patient-reported symptoms which are not validated. The impact of user acquisition strategies is not known. However, the similarity in trends observed across two countries (and for respiratory symptoms, over the same period of time in 2019) adds weight to our findings.

Despite these limitations, the analysis presented in this paper has a number of unique strengths. First, the analysis was conducted using a large, existing dataset that updates in real-time and covers over 1,400 unique self-reported symptoms since November 2016, allowing the monitoring of changes in trends over time. Second, the data is user-driven as a user self-reports their symptoms during an assessment on their own initiative. This allows for identification of changes that would not be detected in traditional studies focusing on specific and/or predefined areas. Third, the large number of symptoms presented in the results that are consistent with expectations, the observation of seasonal differences, and that the results were observed in both Germany and the UK is an indication of the reliability of Ada’s data, demonstrated especially by the detected increase in the less frequent but typical COVID-19 symptoms *hyposmia* and *diminished sense of taste*. While the clinical soundness of Ada’s model at the level of individual diagnostics has already been demonstrated in other studies [54,55], the presented results build confidence that the data collected through the Ada app can also detect health changes in a population in real time.

### Future research

Future research can build on these strengths focusing on reasons for some of the detected changes and by expanding the analysis to more countries. Of particular interest are countries from the Global South and low- and middle-income countries, given the comparative paucity of up-to-date health data in these countries and the differentials in the burden of disease. In addition, investigating changes in trends over time as the implementation of the COVID-19 measures adapts according to the reality in different countries (i.e. as individuals return to work) will offer meaningful insights into the effects of policy changes.

## Conclusions

Our findings suggest that symptom assessment tools might have a role to play in improving understanding of the implications of public health measures. In this analysis we have shown an innovative use of an existing dataset that enables policy makers to inform and monitor public health measures with a real time, low-resource syndromic surveillance system that is relevant both during the COVID-19 pandemic but also in the future.

## Data Availability

When possible, data relevant to the study are included in the article or uploaded as supplementary information.

## Author Statement

AM & FB contributed to the planning (study conception, protocol development) which was revised by CC and AG. AM & FB contributed to the conduct (data collection and data analysis). AM, FB, CC, & AG contributed to the reporting (report writing). All the authors contributed to commenting on drafts of the report. AM is the guarantor for this work. The corresponding author attests that all listed authors meet authorship criteria and that no others meeting the criteria have been omitted.

## Disclaimer

None

## Acknowledgements

Stephen Gilbert (Director of Clinical Evaluation, Ada Health GmbH), Vincent Jörres (Communications & Public Affairs Lead, Ada Health GmbH), Vanessa Lemarié (Director of Business Development & Program Owner of Rare Disease Initiative in Business Development), Nicola Vona (Director of Reasoning Technology in Product), and Paul Wicks reviewed the manuscript and provided feedback.

## Abbreviations

°C: degrees Celsius
COVID-19: Coronavirus disease 2019
GDPR: General Data Protection Regulation
ICD-10: International Classification of Diseases version 2019
Log2 FC: Log2 Fold Change
mm: millimeters
*P*-value: probability value
UK: United Kingdom

